# Potential Lives Saved Through Widespread Global Availability of GLP-1 Receptor Agonists: A Modeling Study

**DOI:** 10.1101/2024.11.11.24317112

**Authors:** David Brook, Peter A. Singer

## Abstract

This model attempts to quantify the potential impact of GLP-1 receptor agonists (GLP-1) in reducing global mortality linked to obesity, type 2 diabetes mellitus (T2DM), and cardiovascular disease (CVD). Using global population data, T2DM and obesity prevalence, cardiovascular risk, and mortality reduction from GLP-1s, we estimate that between ~2.1 million (for people with T2DM or obesity with CVD) and ~3.1 million (for people with T2DM or obesity with/without CVD) lives could be saved annually with widespread global access to GLP-1 receptor agonists.

## Introduction

Obesity and T2DM are major risk factors for cardiovascular disease, the leading cause of mortality globally. GLP-1 receptor agonists, like semaglutide and tirzepatide, have been shown to effectively reduce body weight and improve glycemic control. GLP-1 receptor agonists have also been shown to reduce all-cause mortality (1, 2).

Pandey et al estimated the number of lives that could be saved by expanded access to weight loss drugs in the United States (3). Health equity advocates have also highlighted access disparities in obesity treatments for African Americans (4). However, to our knowledge, there is no estimate of the number of lives that could potentially be saved globally if GLP-1s were more broadly available, nor an advocacy movement for global equitable access.

In this study, we developed a model to estimate the number of lives that could be saved globally through the widespread and equitable availability of GLP-1s. In developing this model, we drew on the most recent peer-reviewed sources to estimate the prevalence and impact on mortality of obesity, T2DM, and cardiovascular disease, noting the disparate geographic scope, timespan, and populations of these studies. We estimated lives saved. Our intention in building this model is not to provide a precise measure of potential benefits, but rather to better understand their size. We also wanted to publicly post a simple and transparent model that others could improve and update as new data becomes available.

This version of our preprint updates our previous publication on November 11, 2024. Compared to the earlier version, a study has shown a higher relative risk reduction in patients with all three of obesity, T2DM, and CVD (5). A new meta-analysis has shown lives saved in patients with T2DM with or without cardiovascular disease (6). Another meta-analysis has found a decrease in all-cause mortality in patients with obesity, but not T2DM, with or without CVD (7).

## Methods

### Model Description

Using Microsoft Excel, we constructed a model to estimate annual lives saved by:

1. Calculating the global population over the age of 19.
2. Estimating the prevalence of obesity, T2DM, and cardiovascular disease in this population.
3. Estimating the impact on mortality of obesity, T2DM, and cardiovascular disease, and combinations thereof.
4. Applying reductions in all-cause mortality associated with the use of GLP-1s.
5. Aggregating potential mortality reduction across global populations with obesity and/or T2DM and cardiovascular disease.

### Data Sources

All data sources are referenced in the model (see **Annex 1**).

- **Global Population:** Based on estimates from the United Nations.
- **Prevalence of Obesity and Diabetes:** Data from the International Diabetes Federation and other public sources were used to estimate global obesity and diabetes rates among adult populations.
- **CVD Risk:** Cardiovascular disease prevalence among adult populations, in general, and among the population of obese adults and adults with T2DM. The associated mortality rates were integrated into the model to calculate the reduction in CVD-related deaths.
- **Obesity, Diabetes, and CVD Mortality Impacts:** Data from the BMC Public Health, NCBI-NLM, and other public sources were used to estimate the mortality impacts of obesity, T2DM, and CVD, and permutations of the three, among adult populations.
- **Mortality Reductions from GLP-1**: Publicly available data from BMC Public Health and from the SELECT and FLOW trials of semaglutide were used to estimate mortality reductions from GLP-1.

See **Supplementary Materials** for the complete model, including references and rationale for each of its elements.

## Results

The results of the model are summarized in **Table 1**.

**Table 1.** Potential Lives Saved from the Widespread Global Availability of GLP-1 Receptor Agonists.

| CO-MORBIDITIES | GLOBAL POPULATION<br>> 19 | ESTIMATED<br>MORTALITY RATE | REDUCTION in<br>MORTALITY from GLP-1 | EXPECTED<br>DEATHS | EXPECTED DEATHS<br>w GLP-1 | LIVES SAVED |
| --- | --- | --- | --- | --- | --- | --- |
| Population > 19 + T2DM + Obesity + CVD | 118,093,278 | 2.6% | 30% | 3,078,692 | 2,155,084 | 923,608 |
| Population > 19 + T2DM - Obesity + CVD | 69,953,343 | 1.3% | 12% | 917,368 | 807,284 | 110,084 |
| Population > 19 - T2DM + Obesity + CVD | 115,656,352 | 1.6% | 19% | 1,821,207 | 1,475,178 | 346,029 |
| Population > 19 + T2DM + Obesity - CVD | 248,656,032 | 1.6% | 12% | 3,928,765 | 3,457,313 | 471,452 |
| Population > 19 + T2DM - Obesity - CVD | 147,293,063 | 1.2% | 12% | 1,750,659 | 1,540,580 | 210,079 |
| Population > 19 - T2DM + Obesity - CVD | 395,594,338 | 1.5% | 18% | 5,847,092 | 4,794,616 | 1,052,477 |
| Population > 19 - T2DM - Obesity + CVD | 308,317,027 | 1.3% | 19% | 4,018,912 | 3,255,319 | 763,593 |
| **All World IMPACT** | **LIVES SAVED** |  |  |  |  |  |
| Any +T2DM and (-T2DM+Obesity+CVD) | 2,061,252 |  |  |  |  |  |
| with +Obesity -CVD -T2DM | 3,113,729 |  |  |  |  |  |
| with +CVD -Obesity -T2DM | 3,877,322 |  |  |  |  |  |

The model estimates that if GLP-1 Receptor Agonists were made available globally, between ~2.1 million (for people with T2DM or obesity with CVD) and ~3.1 million (for people with T2DM or obesity with/without CVD) lives could be saved annually in populations with obesity, and/or T2DM, and/or cardiovascular disease. An additional ~760,000 lives could be saved in patients with CVD alone (without T2DM or obesity). Still, the evidence for the effectiveness of GLP-1s in populations with CVD alone is not yet sufficiently robust to include it in Total Lives Saved.

## Discussion

Our model estimates that widespread global availability of GLP-1 receptor agonists could save between ~2.1 million (for people with T2DM or obesity with CVD) and ~3.1 million (for people with T2DM or obesity with/without CVD) lives annually. This figure underscores the profound public health impact these drugs could have, particularly in reducing all-cause mortality associated with cardiovascular disease (CVD) deaths, obesity, and T2DM. Currently, however, the benefits of GLP-1s are concentrated in high-income countries, with limited access in low- and middle-income countries (LMICs).

This estimate of lives saved from GLP-1s is comparable to the lives saved from childhood vaccines of 154 million lives over 50 years (3 million per year), and more than for HIV drugs of 16.5 million lives over 20 years (0.825 million per year) (8, 9). As such, equitable access to GLP-1s represents an opportunity to save lives on a large scale, emphasizing the need for urgent action to expand access to these treatments.

Countries with the most potential for saving lives are populous countries with high numbers of people with diabetes and obesity, including China, India, the US, Pakistan, and Brazil (10, 11). Those with the greatest potential for reducing prevalence are Pacific small island developing states, where obesity prevalence can reach up to 70% (11). Focusing on both is essential if we want to achieve equity among countries.

From a public health policy perspective, our model also shows that targeting those with both T2DM and obesity could be impactful, because of their high baseline event rates and significant potential reductions in all-cause mortality.

While our model provides a useful estimate of the potential impact of GLP-1s on all-cause mortality in target populations, it has limitations. The mortality impacts of obesity, T2DM, and CVD are estimated based on the best available peer-reviewed evidence, but further refinement will be needed as more data become available. Additionally, rates of reduction in mortality are based on publicly available data from a limited number of trials, which do not provide data on their impact on all combinations of co-morbidities. All-cause mortality was a secondary outcome in many of these trials. Finally, the distribution of imputed risk in the population may not precisely match the participants in the clinical trials. This is a global model, and a country-by-country analysis may provide greater precision and added insights.

We used conservative figures in our estimation of mortality benefit. For example, a retrospective cohort study of the use of GLP1s in Taiwan showed a reduction in all-cause mortality of 52% (12). We used more conservative figures.

Newer GLP-1s such as tirzepatide, often in combination with GIP and Glucagon receptor effects, have been shown to have an even more significant impact on weight loss, although whether this leads to greater reductions in all-cause mortality is currently unknown. Moreover, GLP-1s are also being studied (and show preliminary but promising results) in the treatment of a broader range of conditions beyond CVD. (13)

Our goal was not to provide a comprehensive model of reducing mortality; that has been done in a recent analysis (14). From a policy perspective, some will argue that existing interventions such as taxes on sugar-sweetened beverages, front-of-package labelling, and regulation of ultra-processed foods are more acceptable and/or cost-effective approaches to reduce mortality from obesity, T2DM, and CVD in LMICs. There is evidence that these approaches are effective, but in many countries, they are already being used, and obesity and T2DM rates continue to rise. This suggests the need for further interventions to curb and, ideally, reverse these rates.

Others will argue that the availability of less expensive cardiovascular drugs, such as anti-hypertensives, should be prioritised instead of GLP-1s. Since the mechanisms of all these treatments are independent, and since currently available approaches are not stemming the rise in obesity and T2DM (and presumably the corresponding mortality) in most countries, we suggest that the broader and more equitable GLP-1 access should also be pursued. Indeed, there is early evidence that in the US, the introduction of GLP1s has led to “peak obesity,” reversing this trend for the first time (15).

Modelling the effects of widespread global access is not the same as providing it. Multiple barriers to access would need to be overcome, including manufacturing, affordability, finance, regulatory, and intellectual property. While beyond the scope of this paper, we note the dynamic aspects of this market, including the expiry of the patent on semaglutide in India and China in 2026 (16, 17), with domestic firms preparing to manufacture it.

The precautionary principle suggests that, even in the face of uncertainty, immediate action should be taken to expand access to GLP-1s. Delaying broader access to these drugs could lead to the unnecessary loss of up to a million lives each year. While future trials and research may refine our understanding of these drugs’ impact and, hence, their most impactful deployment, the urgency to act now is clear.

## Conclusion

Expanding global access to GLP-1 receptor agonists offers a unique opportunity to reduce mortality associated with obesity and T2DM. Our model, which estimates that between ~2.1 million (for people with T2DM or obesity with CVD) and ~3.1 million (for people with T2DM or obesity with/without CVD) lives are lost annually, highlights the critical importance of overcoming barriers to access. Through innovative financing, advocacy, and global cooperation, the world can seize this opportunity to save lives on an almost unprecedented scale.

## Supporting information

Supplemental Model 1

## Data Availability

Model is provided in the Supplementary Materials. All data produced are available in the references cited in the model.

## Acknowledgments

Thank you to colleagues at World Obesity Federation, Ole Frithjof Norheim, Haidong Wang, and another reviewer for feedback and guidance on data sources and on the model, and to Jocalyn Clark for reviewing an earlier version of this paper.

## Disclosures

David Brook worked part-time with Gilbert’s Solutions (<u>gilbertslaw.ca</u>) when the first version of this model was created; Gilbert’s Solutions has received advisory fees from Novo Nordisk but not for the development of this global model. Peter Singer owns shares in GLP-1 companies.

An initial draft of this paper was developed with the assistance of ChatGPT-4.

